# Experiences of supported isolation in returning travellers during the early COVID-19 response: an interview study

**DOI:** 10.1101/2021.02.17.21251735

**Authors:** H. Carter, D. Weston, N. Greenberg, I. Oliver, C. Robin, G. J. Rubin, S. Wessely, R. Amlôt

## Abstract

**Objectives:** To understand the experiences of those who underwent supported isolation as part of the response to the COVID-19 pandemic, after returning to the UK from Wuhan, China.

**Design:** We used semi-structured interviews to capture participants’ experiences and perceptions of supported isolation.

**Setting:** Telephone interviews carried out within approximately one month of an individual leaving supported isolation.

**Participants:** 26 people who underwent supported isolation at either Arrowe Park Hospital (n = 18) or Kents Hill Park Conference Centre (n = 8) after being repatriated from Wuhan in January – February 2020.

**Results:** Participants were willing to undergo supported isolation because they understood that it would protect themselves and others. Positive treatment by staff was fundamental to participants’ willingness to comply with isolation procedures. Despite the high level of compliance, participants expressed some uncertainty about what the process would involve.

**Conclusions:** As hotel quarantine is introduced across the UK for international arrivals, our findings suggest that those in charge should: communicate effectively before, during and after quarantine, emphasising why quarantine is important and how it will protect others; avoid enforcement and focus on supporting and promoting voluntary compliance; facilitate shared social experiences for those in quarantine; and ensure all necessary supplies are provided. Doing so will increase adherence and reduce any negative effects on wellbeing.

## Introduction

The first cases of a novel strain of coronavirus (SARS-CoV-2) were detected in Wuhan, China, in December 2019. On 31^st^ January 2020, British Nationals living in Wuhan were offered repatriation to the UK. 93 returned on two chartered flights. In order to be repatriated all had to agree to undergo 14 days of ‘supported isolation’ (i.e. quarantine). This took place in an accommodation block at Arrowe Park Hospital in the Wirral [1]. A further 118 people returning from Wuhan underwent supported isolation at Kents Hill Park Conference Centre, Milton Keynes. All supported isolation ended by 23^rd^ February 2020 [2].

Supported isolation for returning travellers had, to our knowledge, never been used before within the UK. It was anticipated that the experience could have considerable psychological consequences for the individuals concerned, including potential post-traumatic stress, anger and confusion; consequences that may be affected by a range of stressors including information provision, stigma, and fear of infection [3]. Furthermore, supported isolation represents a unique social context in which relative strangers are placed in close quarters within a novel context and asked to adhere to recommended behaviours for a prolonged period. During emergencies, such social contexts can affect individuals’ social identity, which can have consequences for adherence and psychological resilience [4,5,6]. Outside of the emergency response context, the emergence of strong social connections among strangers in close physical proximity has been associated with positive well-being related outcomes [7].

From 15^th^ February 2021 those travelling to the UK from some other countries will be required to isolate in hotels for 10 days [8]. Policy around this isolation is focused on identifying the best ways to maximise compliance, with an increasing emphasis on enforcement [9]. Furthermore, with the COVID-19 pandemic ongoing, it is possible that supported isolation will be required in other contexts, such as to assist those with difficulty isolating at home [10] and to reduce household transmission [11]. It is therefore important to understand more about the way in which people experience supported isolation, so that this process can be optimised to increase adherence and mitigate any negative effects on wellbeing. We carried out a rapid mixed-methods study in which we interviewed individuals who underwent supported isolation at Arrowe Park Hospital and Kents Hill Park conference centre. To our knowledge, this is the first research conducted with individuals during and immediately following their supported isolation in this country. These experiences are once again topical in light of the upcoming policy on required hotel isolation.

## Method

### Ethical approval

Ethical approval was obtained from the Public Health England Research Ethics Governance Group (approval no. NR0187).

### Patient and public involvement

Given the extremely rapid and responsive nature of this research, it was not possible to involve patients or the public in the development of the study and associated materials. However, staff at the supported isolation facilities were involved from the outset in planning the study and facilitating participant recruitment. Additionally, findings from this study will be shared with participants on publication.

### Design

This study used semi-structured interviews to capture participants’ experiences and perceptions of supported isolation. Interviews took place over the telephone, within one-month post-supported isolation. The study was designed and carried out in-line with consolidated criteria for reporting qualitative research (COREQ) guidelines [12] (see Appendix 1).

### Participants

Participants underwent supported isolation in either Arrowe Park (n = 18) or Kents Hill Park (n = 8) in January and February 2020. The day before leaving supported isolation, participants were provided with an information sheet about the study by a member of staff at the facility. This included an invitation to take part in a survey (findings reported elsewhere), as well as the opportunity to take part in an interview. On leaving supported isolation, 69 people provided a contact email address, and all were invited to take part in an interview. Of these, 26 people (38%) consented to take part in an interview, this sample therefore represents 12.3% of the entire population who underwent supported isolation. Half of the participants (n = 13) were male and half (n = 13) were female. Participants ranged in age from 22 to 78.

### Materials

An interview schedule was developed to capture in-depth information about individuals’ experiences and perceptions of supported isolation, including their: overall experience (e.g. “Tell me about your experience of undergoing supported isolation”); willingness to undergo supported isolation (e.g. “Were you willing to undergo supported isolation”); perceptions of the way the supported isolation process was managed (e.g. “In general, how do you feel the supported isolation process was managed?”); perceptions of others’ behaviour during supported isolation (e.g. “How did those in supported isolation behave towards each other?”); experiences after leaving supported isolation (e.g. “How has life been for you since leaving supported isolation?”).

### Procedure

Each interview took place within one month of leaving the supported isolation facility and lasted for approximately an hour. Interviews were carried out by behavioural scientists based at Public Health England or King’s College London, all of whom were qualified to at least MSc level and had received training in carrying out interviews. Researchers did not establish a relationship with participants prior to carrying out the interview nor were participants made aware of any personal characteristics of the interviewer, aside from their place of work and the broad aims of the research. Interviews were carried out by both male and female members of the research team. Only the researcher and the participant were present during the interview. Interviews were recorded and subsequently transcribed. After taking part in an interview, participants received a debriefing statement which provided further information about the study, as well as sources of support that participants could access if required. Participants were informed that they could request a copy of the results but did not provide feedback on the findings.

### Analysis

A framework approach was used to analyse the data. This is a type of thematic analysis that is commonly used within research that has implications for policy and practice [13]. An a priori thematic framework was developed, but themes were also allowed to emerge from the data. This analysis generated 12 key themes: compliance; feelings about undergoing supported isolation; risk perceptions around catching COVID-19; protective behaviours during supported isolation; management of supported isolation; treatment by staff and authorities; communication from staff; communication with those outside of supported isolation facilities; relationship with others within supported isolation; thoughts about others’ behaviour during supported isolation; areas for improvement; feelings on leaving supported isolation. Analysis was carried out by hand by the first author, and each passage was coded into one or more of the identified themes. After analysing the 26 transcripts no new themes emerged, thus data saturation had been reached [14].

## Results

Results are presented by theme below; supporting quotes are presented in Appendix 2.

### Compliance

Most participants were willing to undergo supported isolation. They understood why supported isolation was necessary and why they were being asked to undergo it. In the few instances where participants did not want to comply, non-compliance took the form of breaking the rules inside the supported isolation facility (e.g. trying to obtain more alcohol than was allowed), but not trying to leave the supported isolation facility.

### Feelings about undergoing supported isolation

As well as discussing their willingness to comply with supported isolation measures, participants also discussed their feelings about undergoing supported isolation more broadly.

Most participants felt that the positives of supported isolation outweighed the negatives. Positive aspects were grouped broadly into three themes: a belief that supported isolation protects family and friends as well as UK society; a belief that supported isolation would protect themselves, by ensuring they were in a safe place if they developed symptoms and that they would not be blamed in the event of an outbreak in the UK; and faith in the effective management of the supported isolation process.

Where participants expressed concerns these centred around uncertainty about what the process would involve, sometimes attributing this to lack of information being provided. Others were concerned that they would be bored or would be at increased risk of catching COVID-19. A few felt angry or frustrated about the process, because they didn’t think it was necessary or believed it was a waste of time and resources.

### Risk perceptions around catching COVID-19

Participants’ reported different perceived risks of catching COVID-19 whilst in isolation. Some felt at low risk because they could take protective behaviours; that anyone displaying symptoms could be quickly isolated; and that everyone in the supported isolation facility underwent regular testing.

However, others were very worried due to other people having symptoms, and the need to sometimes be in close proximity to others. In general, most participants stated that their risk perception reduced over time in the facility, as people continued to test negative, and did not have any symptoms. The majority of participants noted that they felt most worried at the start of supported isolation process.

### Protective behaviours during supported isolation

The majority of participants reported that they performed protective behaviours. The most common included staying in their own room, observing effective hand hygiene measures, and wearing a face mask. While most participants reported that they took at least some protective measures, those who took fewer measures often reported that this was due to their perception that the risk of catching COVID-19 during supported isolation was low.

### Management of supported isolation

Most participants reported that they felt the whole process was well-managed. Reasons for this included that the process was well-organised, and that staff and management were willing to adapt procedures following negative feedback about the process.

Where participants did express concerns these often centred on provision of food, for example not receiving meals, poor food options, food being served uncovered, and food not being warm enough. Another area of management that participants suggested could be improved was around internal communication within and between organisations. For the most part, participants who provided negative feedback about the management of the supported isolation process felt that changes were made to address their concerns, and that the management of the supported isolation process improved as time went on.

### Treatment by supported isolation staff and authorities

Overall, participants were extremely positive in their feedback about the way in which staff treated them. The staff were friendly and helpful, went out of their way to keep people happy, and provided people with anything that they asked for. A few participants mentioned that staff did not try to avoid them or treat them as if they were ill. A small number of participants specifically noted that staff achieved a good balance between promoting good public health, without making the process too restrictive.

### Communication from staff during supported isolation

Participants were also overwhelmingly positive about the way in which members of staff communicated with them. Almost all participants talked about the daily newsletter that they received from staff and felt that this was an effective way of providing information about protective actions, timings of any activities, and testing. Similarly, participants noted that staff were proactive in their communications, calling regularly to check on each individual, and scheduling regular update meetings. Participants also felt that staff answered all their questions (or tried to) and were open and transparent in providing information.

Some expressed dissatisfaction at the somewhat old-fashioned methods of communication, inability of staff to answer some questions, and information not being provided in multiple languages.

### Communication with those outside of supported isolation facilities

Most participants found it easy to communicate with those outside supported isolation and did so regularly. Several participants expressed how important this was in helping them to get through the supported isolation process. Additionally, some were able to carry on working during supported isolation, and this helped them to pass the time. A few participants also highlighted the benefit of local community groups who posted pictures of uplifting things.

On the other hand, some participants did note difficulties in communicating with those outside of supported isolation, and these typically related to having limited access to internet or poor phone signal.

### Relationship with others within supported isolation

Where people felt a connection with others this was often due to a sense of camaraderie or shared experience. Some participants described how people supported and encouraged each other during the supported isolation process, stating that this helped people to get through the experience. This connection was facilitated by the formation of chat groups, and some level of freedom to socialise with others.

Where people did not feel a connection with others this was because they either didn’t get the opportunity to interact much with others, or actively avoided it (due to fears about catching COVID-19).

### Thoughts about others’ behaviour during supported isolation

Most participants felt that they could trust others to behave appropriately and instances of uncooperative behaviour were rare or non-existent. A handful of participants noted isolated instances of uncooperative behaviour, but almost all said that the majority of people were friendly and cooperative.

### Areas for improvement in supported isolation procedures

Some felt they would have liked more information about what supported isolation would involve. Others suggested it would have been beneficial to have more access to outside space and exercise facilities. Another common area for improvement was the food provided, with people suggesting that food options and quality could have been better.

### Feelings after leaving supported isolation

Many participants felt happy and relieved to leave supported isolation and get back to normal. However, several participants stated that they struggled after leaving supported isolation. Some felt anxious or overwhelmed, with reasons including not being used to going outside, or being concerned about mixing with large numbers of people again. Others simply stated that they had generally struggled on leaving, or that they had experienced negative reactions from others.

The majority of participants did not receive follow up information, though a few did receive information about sources of further support. While some stated that they would not have expected to receive any additional information, others felt that this would have been helpful.

## Discussion

This paper represents the first in-depth analysis of the experiences of those who underwent supported isolation in the UK during the first wave of the COVID-19 pandemic. Given that supported isolation is once again required in the management of COVID-19 [8,10,11], our findings should help facilitate optimised management.

Despite some initial concerns, including confusion about what the process would involve and fears of infection, all willingly complied with the voluntary supported isolation process. People understood why it was necessary and believed that doing so would protect themselves, their friends and family, and others in the UK; motivation for adherence was largely altruistic. Participants were overwhelmingly positive about their treatment by staff, communication from staff, and overall management of the supported isolation process. This was fundamental to participants’ willingness to comply with the restrictions of their liberty. Our findings are in line with systematic reviews carried out at the start of the pandemic [3,15], as well as research into the management of other types of emergencies [4,16]. Crucially, participants believed their treatment by staff was legitimate, and they therefore chose to comply with supported isolation procedures; it is likely that compliance would have been much lower had staff attempted to enforce compliance [16].

There were mixed views as to whether people in isolation experienced a connection with each other. However, almost all reported that others were helpful and friendly. Additionally, a number of people developed a shared identity with others; for example, they talked about everyone being in it together or going through the same experience. Those that did develop a shared identity often reported that this helped them to get through the process. This is as would be expected based on previous research which suggests that when people experience a sense of shared identity with others, this promotes adherence to protective measures, resilience and well-being [4,5,7]. While a sense of shared social identity arose spontaneously in some instances, participants emphasised that being able to communicate with others (for example, via chat groups) enhanced the social support that they experienced. Promoting virtual interaction between those undergoing supported isolation may be beneficial for strengthening shared identity, facilitating provision of social support, and promoting resilience and well-being. Participants also highlighted how important it was that they were able to easily keep in touch with friends and family during the supported isolation process. Of particular interest was our finding that some participants reported negative experiences on leaving supported isolation. It may therefore be beneficial to prepare participants for possible psychosocial reactions prior to them leaving supported isolation and signpost them to sources of support.

### Limitations

Approximately a third of participants who were contacted about this study agreed to take part. We have no information on those who did not participate, and it is possible that they differed on key variables. Of those that did participate we reached thematic saturation within the sample. Furthermore, participants were aware of what was going on around theme, so the reports of very high compliance with supported isolation and other protective behaviours can be generalised to all those who were in quarantine. The same goes for the general finding that most people were friendly and cooperative. A second limitation is that only those who had a good understanding of English were interviewed. It is therefore possible that the experience differed for those who were less able to understand English; indeed, this was alluded to in some comments made by participants. A final limitation is that this study was jointly run by King’s College London and Public Health England, and Public Health England also assisted with the management of the supported isolation process. Although the team carrying out this research were not associated with the management of the supported isolation process, it is possible that participants were aware that PHE played a role in managing the supported isolation process, and that this affected their responses during the interview.

## Conclusion and Recommendations

The findings presented here, particularly when situated within the wider literature, generate several key recommendations that are particularly relevant given the upcoming requirement for travellers to isolate in hotels. The supported isolation carried out in January – February 2020 was designed to support those who were returning to the UK, and every effort was made to ensure that their experience was as positive as possible; as participants noted, staff could not do enough for them. Isolation in hotels is likely to be fundamentally different, with limited support from staff and an emphasis increasingly on enforcement rather than encouragement [9]. The reasons why people are travelling in the middle of a pandemic will also be different. The UK may find itself placing people into isolation who are more likely to experience distress such as those who are arriving to attend a funeral, are travelling due to a family crisis, or who do not speak English. We must also not forget that, unlike travellers placed into facilities at Arrowe Park or Kents Hill, returning travellers will now be asked to pay £1,500 each towards their isolation. It is therefore critical that those responsible for implementing policies on isolation requirements take into account the recommendations presented here; failure to do so is likely to reduce adherence to isolation and risks serious long-term impact on those involved.

Specific recommendations are: 1) prior to supported isolation, communicate with those affected about why isolation is necessary, how it will help to protect others, and what the process will involve. Given that compliance is often motivated by altruism, emphasising how isolation will protect others is crucial. Such communication will also reduce concerns related to uncertainty about the isolation process; 2) communicate effectively with those undergoing isolation, throughout the process. Communication should be open and honest, and information should include protective actions people should take, why taking such actions is effective, and how taking such actions protects oneself and others; 3) enforcement with isolation should be avoided wherever possible. Given the large numbers of people who may be required to isolate at one time it will not be possible to enforce adherence; attempting to do so is likely to be perceived as illegitimate, thereby reducing adherence and risking serious long term consequences for those involved; 4) facilitate and encourage development of shared identity among those undergoing supported isolation, via the formation of chat groups or other means of communication, that include staff managing the facilities. This type of shared social identity should encourage both adherence to supported isolation measures, and improved resilience during the supported isolation process; 5) ensure that all essential supplies (such as food, exercise facilities, ability to communicate with those outside isolation) are provided and are suitable for the needs of the traveller; 6) provide information prior to leaving supported isolation to help people to prepare to return to their normal lives. This should include information about emotions that people might experience, and sources of further support that people can access if required. It may also be beneficial to include in this information any ongoing expectations around adherence to protective behaviours.

## Data Availability

Following peer review, data will be available on request from the corresponding author.

## Competing interest statement

All authors have completed the ICMJE uniform disclosure form and declare: HC, DW, IO, CR, and RA are current employees of Public Health England; GJR participates in the UK’s Scientific Advisory Group for Emergencies and its subgroups.

## Transparency declaration

The lead author affirms that this manuscript is an honest, accurate, and transparent account of the study being reported; that no important aspects of the study have been omitted; and that any discrepancies from the study as originally planned (and, if relevant, registered) have been explained.

## Contributor statement

RA, HC, DW, NG, GJR, IO and SW conceived the study. HC, CR, DW and RA collected the data. HC carried out the analysis and wrote the first draft of the manuscript. All authors contributed to the design and implementation of the study, and to the writing of the manuscript.

## Data sharing statement

Data are available on request.

## Funding statement

HC, DW, GJR, NG, RA and SW are supported by the National Institute for Health Research Health Protection Research Unit (NIHR HPRU) in Emergency Preparedness and Response, a partnership between Public Health England, King’s College London and the University of East Anglia. DW, IO, CR and RA are supported by the NIHR HPRU in Behavioural Science and Evaluation, a partnership between Public Health England and the University of Bristol. CR is also supported by the NIHR HPRU in Emerging and Zoonotic Infections and NIHR HPRU in Gastrointestinal Infections. The views expressed are those of the authors and not necessarily those of the NIHR, Public Health England or the Department of Health and Social Care.

