## Appendix 2 for "Experiences of supported isolation in returning travellers during the early COVID-19 response: an interview study"

**Appendix 2: Supporting quotes**

| **Sub-theme** | **Supporting quotes** |
| --- | --- |
| **Compliance** | |
| Understanding why supported isolation is necessary | *“I understood the necessity and I was willing to cooperate very much”* (P2)  *“I was prepared to [undergo supported isolation] […] I knew it was a necessary evil”* (P19) |
| Non-compliance through rule-breaking | *“Over a short period of time it was let’s try and break the rules just for something to do. Let’s see how far we can go”* (P4) |
| **Feelings about undergoing supported isolation** | |
| ***Positive feelings about supported isolation*** | |
| Supported isolation protects others | *“we were a risk to others and we didn’t want to come back to our families carrying the virus”* (P18)  *“it was in our best interests and the people we love in the UK and the country in general”* (P8) |
| Supported isolation protects self | *“in the event that I or any of my fellow travellers developed symptoms we would be in that hospital environment or we would be with doctors who spoke our native language”* (P9)  *“[supported isolation is important] because I would have felt very bad if it had have been found out later that it was from myself that had passed on or started something”* (P11) |
| Faith in effective management | *“when we actually arrived at Arrowe Park […] the staff there gave such a warm welcome and made everything feel so sort of warm and comfortable”* (P16) |
| ***Negative feelings about supported isolation*** | |
| Uncertainty about supported isolation | *“You’re thinking well what are the facilities here going to be like? How am I going to cope with that?”* (P24)  *“I was a little bit apprehensive just because I didn’t know […] how it would be structured or organised, and obviously the lack of details”* (P11) |
| Boredom | *“[I was concerned that] I would be a bit bored”* (P19) |
| Increased risk of catching COVID-19 | *“Our biggest concern would be is anybody sick because of this virus among us?”* (P2) |
| Thought supported isolation unnecessary | *“we did think it was unnecessary because we were already tested negative”* (P24) |
| Believed supported isolation a waste of time | *“it was an over the top response that probably cost 2 or 3 million pounds for those two weeks”* (P4) |
| **Risk perceptions of catching COVID-19** | |
| ***Low perceived risk*** | |
| Could take protective behaviours | *“we were just very careful with washing our hands […] just sensible hygiene precautions really. So that made us feel pretty safe”* (P16) |
| Rapid isolation of those with symptoms | *“I knew that things were being monitored very carefully and things were being done about it”* (P4) |
| Regular testing | *“after one week we’d all been tested negative, after 10 days we’d all been tested negative, after 14 days we’d all been tested negative”* (P4) |
| ***High perceived risk*** | |
| Others had symptoms | *“someone with a high temperature, she was really close to me, so I said oh please don’t stay too close”* (P10) |
| Close proximity to others | *“we were using the same big meeting room for one or two hours before we eventually went to our separate rooms”* (P2) |
| Reduced risk perception | *“after the first week we didn’t see anyone showing symptoms so everyone was more relaxed talking to each other”* (P10) |
| **Protective behaviours** | |
| Stayed in own room | *“we just decided not to go out, just to stay in our hotel rooms”* (P2) |
| Hand hygiene | *“I would wash my hands when I went downstairs”* (P11) |
| Wearing a face mask | *“we were wearing gloves and masks and keeping no more contact with each other”* (P3) |
| **Management of supported isolation** | |
| ***Process was well managed*** | |
| Process was well-organised | *“the place all sort of ran like clockwork from my point of view”* (P16)  *“I don’t think they could have done it any better”* (P21) |
| Process was adapted following feedback | *“the food initially it was only microwave meals but that evolved in the second week […] everybody was sort of learning as we went along”* (P5)  *“after the first 4 or 5 days everything was up and running and was very smooth and very supportive”* (P12)  *“they are improving their responding and they are learning from their mistakes as well they were really good I was really impressed”* (P6) |
| ***Process was not well managed*** | |
| Meals not received | *“they forgot to give me breakfast and lunch three times”* (P1) |
| Poor food options | *“when they chose a facility that didn’t have fresh food on site they didn’t understand the Chinese way of life”* (P8) |
| Food served uncovered | *“I think most of us had the salad or the bread which was not covered”* (P2) |
| Food not warm enough | *“the food turned up lukewarm in cardboard boxes”* (P22) |
| Communication between organisations | *“With the change of shifts, they didn’t update people […] there was no passing on of communication, there was no register of requests from room numbers”* (P18)  *“there was three branches of the NHS staff there and they all had their own rules […] so it’s very difficult to merge as one”* (P4) |
| **Treatment by supported isolation staff** | |
| Staff friendly and helpful | *“we were treated with compassion […] and so we were immediately put at ease”* (P24)  *“the staff there were really helpful and they were lovely”* (P21) |
| Staff tried to keep people happy | *“the staff went above and beyond in trying to help us”* (P11) |
| Staff provided everything people asked for | *“if you needed anything you could get it brought to your room within an hour or two”* (P22)  *“staff were very helpful, whatever we asked they tried to answer, and whatever we needed they tried to procure”* (P13) |
| Staff did not treat people as if they were ill | *“we don’t feel that really we were isolated or we were frightening […] as somebody who might carry a virus”* (P2)  *“they were actually treating us like normal people”* (P7) |
| Process did not feel too restrictive | *“I think that’s a balance that had to be struck between health risk and […] how we felt that we were being treated, how restricted we felt”* (P11) |
| **Communication from staff** | |
| ***Positive aspects of communication*** | |
| Daily newsletter | *“I think they were really good…we would get two or three letters a day actually sometimes about what was changing and why”* (P13) |
| Proactive communication | *“in the mornings when a nurse would come around […] if there were any sorts of developments to tell us about then they would”* (P15) |
| Staff answered all questions | *“I think they would have answered anything that we needed to know”* (P22) |
| ***Negative aspects of communication*** | |
| Staff could not always answer questions | *“the only information [that staff couldn’t give me] was sort of about leaving actually, and what was going to happen […] that information was only very near the end”* (P15) |
| Communication methods old-fashioned | *“their way of disseminating information was posting things under the door, which […] seems a little old-fashioned […] maybe if they had done a group chat or done a group email […] I think that may have been a good way of communicating”* (P16) |
| Information not provided in multiple languages | *“the Mum […] had to ask for a lot of help because of her difficulties with English, she was a Chinese national”* (P11) |
| **Communication with those outside isolation** | |
| Communication with those outside important | *“we spent half the day usually emailing and skyping and WhatsApping everybody […] it was actually good having that routine”* (P24) |
| Benefit of local community groups | *“it’s nice when you are in that situation […] to see stuff that wasn’t about the virus, and wasn’t doom and gloom”* (P25) |
| Difficulty communicating with those outside | *“the phone signal where we were was terrible”* (P22) |
| **Relationship with others** | |
| People felt a connection | *“I think there was a bit of camaraderie […] everyone was in the same situation really”* (P16)  *“we were all in the same boat […] it was just, we were all in it together really”* (P22) |
| People supported and encouraged each other | *“we look after each other, we tried to be helpful with each other as well”* (P3) |
| Others’ support improved the experience | *“we encouraged each other and things like that sometimes. It was good to help many to spend the long and sometimes worrying days”* (P2) *“there was definitely a case of we’re going to get through this together”* (P11) |
| Connection facilitated by chat groups | *“we would message on Facebook and WhatsApp and all that stuff”* (P23)  *“we had a little common room within our side of the conference centre […] so we did movie nights and quizzes and things like that”* (P5) |
| ***People did not feel a connection*** | |
| No opportunity to interact | *“the circumstances didn’t really permit interaction”* (P6) |
| Chose not to interact | *“they all got together and things like that and the invitation was open but at the same time I didn’t really want to be in the same room with lots of people”* (P13) |
| **Perceptions of others’ behaviour** | |
| People trusted others | *“people were very very well-behaved […] people are grateful that was a common feeling”* (P6) |
| Isolated instances of non-cooperation | *“there’s only one argument that we ever heard in the whole two weeks and it was somebody saying that they’ve been tested negative three times can they go home early […] but apart from that the whole two weeks was like with no issue at all”* (P8) |
| Most people friendly | *“Almost all […] were quite cooperative [….] I think they were quite friendly to each other”* (P2) |
| **Areas for improvement** | |
| More information in advance | *“in advance, it would have been nice to have been told information as to how it works, in terms of freedom of movement and the ability to cook and stuff for yourself […] it’s the communication, and setting expectations”* (P25) |
| Access to outside space | *“outdoor space improvements may have been helpful […] I think we are all finding value in still being able to get outside a little bit”* (P12) |
| Access to exercise facilities | *“having a couple of running machines and a couple of bikes and things like that would have been really helpful”* (P5) |
| Improved food | *“there wasn’t enough thought put into the requirements of people that were from China […] they didn’t understand the Chinese way of life which is very much fresh food based”* (P8) |
| **Feelings after leaving supported isolation** | |
| Felt anxious or overwhelmed | *“I actually had a panic attack when I got in the taxi I found everything very overwhelming […] I hadn’t really mentally prepared myself for going outside”* (P23)  *“First time we went to the supermarket […] just seeing people who were not in masks and protective clothing took some getting used to. Just being able to wander around and walk […] and all the crowds of people in the supermarket when we’d just been used to us two was quite uncomfortable”* (P24) |
| Struggled on leaving | *“the last night we were there, there was no sense of jubilation […] it was just very quiet, very subdued. […] [Leaving] affected me quite badly really […] I was absolutely lost”* (P4)  *“I stayed at home but I didn’t do a great deal except sleep and I was not in the best of moods most of the time…or part of the time”* (P13) |
| Negative reaction from others | *“the driver who came to pick us up said ‘I will have to call head office to get the car disinfected after I drop you off’ – that response I think will stay with me for a long time”* (P13) |
| Follow up information would have been helpful | *“I understand there is a lot happening right now…but I don’t think there was enough support for us leaving”* (P23) |
